# Unveiling Genetic Associations: Investigating *CLDN16, GRID2, NRG3,* and *CACNG4* Gene Polymorphisms with Insulin Resistance Risk Among Normal BMI Individuals in the Indian Population

**DOI:** 10.1101/2024.08.22.24312447

**Authors:** Sabitha Thummala, Junaid Ahmed Khan Ghori, Sarah Fathima, Katherine Saikia, Nithya Kruthi, Shanti Lakshmi Duraimani, AR Balamurali, Rahul Ranganathan

## Abstract

Studies estimate that India has about 65+ million diabetic patients with a substantial impending increase, making it the international ‘diabetes capital’. Diabetes Mellitus is a metabolic disorder which is signified by elevated blood sugar levels due to defects in insulin action, secretion or both. Insulin resistance (IR) or insulin resistance-linked obesity is also known to be a causing factor of Metabolic syndrome which is a combination of cardiovascular risk factors that include raised fasting plasma glucose, central obesity, hypertension, raised triglycerides, and reduced High-Density Lipoprotein (HDL) cholesterol. This study investigated the association between four single nucleotide polymorphisms (SNPs) in the selected genes - rs6801387 (*CLDN16*), rs72872727 (*GRID2*), rs1414756 (*NRG3*), and rs8065294 (*CACNG4*) and (IR) among a normal BMI Indian population. Through Chi-Square tests, we detected significant associations between SNP genotypes and (IR). Allele frequency analysis revealed higher frequencies of allele G (rs6801387) and T (rs72872727) among individuals with HOMA2-IR >2, while allele T (rs8065294) indicated decreased risk, emphasizing the relevance of genetic factors in metabolic disorders. The differences in clinical parameters such as fat mass, serum triglycerides and HbA1c between the cases and controls highlights the multifactorial nature of the condition. Inheritance model suggested the dominant inheritance for rs6801387 and rs72872727 and codominant inheritance for rs1414756 and rs806529, offering insights into genetic associations with IR. Despite the study’s moderate sample size,these genetic biomarkers exhibit strong susceptibility to the studied condition, showing the importance of exploring their functional significance and underlying biological mechanisms in future research endeavours.

## INTRODUCTION

Diabetes is amongst the top 10 causes of death worldwide, checking off as the biggest worldwide health catastrophe of this century. The WHO reports that in 2019, 74% of deaths were caused due to noncommunicable diseases, out of which 1.6 million deaths were caused due to diabetes [1]. Further it estimates that 592 million people will succumb to diabetes by 2035 [2]. The burden of diabetes is increasing worldwide especially in emerging nations like India where 77 million people were diagnosed with diabetes in 2019. It is estimated that this number would reach 134 million by 2045, amongst which 57% of the affected population will never be diagnosed [3]. One of the methods to combat this huge growing epidemic is focusing on early diagnosis of Diabetes. This may help in managing and also preventing diabetes complications to quite an extent [4]. It will also help relieve the healthcare and economic burden of diabetes care which is high and rising worldwide [5].

Body Mass Index (BMI) is considered a key measure of metabolic syndrome, but recent studies suggest it may not be the only criteria [6,7]. Instead, body composition, specifically muscle and fat mass, offers a more refined understanding [6,8]. While overweight people are more likely to meet metabolic syndrome criteria, it is suggested that higher BMI isn’t necessarily linked to metabolic syndrome.[9]. Although obesity is a recognized risk element for metabolic disorders, not all obese individuals develop these conditions [10,11,12]. Estimates suggest that between 15% to 45% of obese people are metabolically healthy [13,14] while 6% to 30% of normal-weight individuals exhibit cardiometabolic abnormalities typical of obesity [15,16, 17 18].This trend has been reported in Indian population as well. [17 18] The prevalence and categorization of this unconventional drift varies widely due to the lack of standardized definitions [19, 20, 21]. Usually parameters like body fat percentage and waist circumference are considered along with the WHO’s BMI cutoff [22]. This variability throws light on the complexity of metabolic health and emphasizes the need for more defined and standardized criteria and parameters other than BMI alone [19, 20].

A recent genetic study has established associations between specific loci and body fat percentage, indicating its protective effects on glycaemic and lipid outcomes. Individuals with higher genetic susceptibility to IR tend to have lower overall adiposity, suggesting impaired fat storage may harm metabolism [23].

The aim of this study was to investigate the potential genetic influence of polymorphisms in four particular genes in light of previous findings.-*CLDN16, GRID2, NRG3, and CACNG4* on IR in an Indian population. We have analyze four SNPs within these genes (rs6801387 in *CLDN16* rs72872727 in *GRID2* rs1414756 in *NRG3* rs8065294 in *CACNG4*) to assess their association with IR risk in Indian population group.

## MATERIALS AND METHODS

### 1. Study Participants

The research enrolled 191 participants (90 men and 101 women) of Indian ethnicity residing in India, aged 18-65 years, between January 2022 and December 2023. The Homeostasis Model Assessment Insulin Resistance (HOMA2-IR) was used to categorize participants into cases and controls. Individuals with HOMA2-IR > 2 were classified as cases (57 participants), and those with HOMA2-IR < 2 (134 participants) were categorized as controls. The HOMA2-IR was calculated using the online tool provided by the Medical Science Division of The University of Oxford [24]. Participants’ demographics were collected through an electronic registration questionnaire, including self-reported height (cm), and weight (kg). Body mass index (BMI) was calculated as weight (kg)/height (m^2) [25], Body Fat Percentage (BFP) for adult male was calculated as 1.20 x BMI + 0.23 x Age - 16.2, Body Fat Percentage (BFP) for adult female was calculated as 1.20 x BMI + 0.23 x Age - 5.4 [26], and Fat Mass was calculated as BFP x Weight x 0.01 [26]. To ensure participant safety and ethical consideration, the study included only individuals free from: cancer history, cardiovascular or renal failure, mental illnesses, and pregnancy or lactation. All participants provided written informed consent following the principles outlined in the revised Declaration of Helsinki (2008) [27]. The study protocol was approved from the Answer Genomics Ethical Review Committee.

### 2. Laboratory Measurements

Following a 12-hour fast, blood samples were obtained from participants to assess various metabolic markers, such as fasting plasma glucose (FPG), total cholesterol (TC), triglycerides (TG), high-density lipoprotein cholesterol (HDL-C), low-density lipoprotein cholesterol (LDL-C), and glycated haemoglobin (HbA1c). The assessments were conducted using Beckman DxC 700 AU for all markers except HbA1c, which was analyzed with Tosoh G-8, and fasting insulin levels measured by Beckman UniCel DxI 800, adhering to the protocols provided by the manufacturers. The reference ranges for these markers were established as follows: FPG (70-100 mg/dl), TC (0-100 mg/dl), TG (0-150 mg/dl), HDL-C (40-60 mg/dl), and LDL-C (0-100 mg/dl). To determine insulin sensitivity, the Homeostasis Model Assessment of Insulin Resistance (HOMA2-IR) was applied, calculated from fasting plasma glucose (in mmol/l) and fasting insulin (in mU/l) using the formula: fasting glucose x fasting insulin (22.5). Participants with a HOMA2-IR score above 2 were categorized as insulin resistant, while those with scores below 2 were considered insulin sensitive. The clinical characteristics of the study subjects and the distribution of these groupings are detailed in Table 2.

### 3. Genotyping and Single-nucleotide polymorphism(SNP) selection

This study investigated potential genetic influences on IR by analyzing participants’ DNA. The extraction of DNA was conducted on blood specimens utilizing the Qiagen blood extraction kit, acclaimed for its capability to isolate genomic DNA of superior quality from blood samples. The process of genotyping (SNPs) was executed with the aid of the Illumina Infinium Global Screening Array (GSA) V3 platform, recognized for its extensive representation of genetic variance throughout the genome [28]. The Illumina iScan system was used for genotyping, while Genome Studio V2 software was used for interpreting the raw intensity data, quality control, and data export [29].

The study focused on four genes (*CLDN16*, *GRID2*, *NRG3*, and *CACNG4*) involved in metabolism and potentially influencing insulin resistance, as indicated in Table 1. The selection of SNPs was based on a comprehensive scientific literature identifying genetic variants influencing metabolic traits.

**Table 1.**
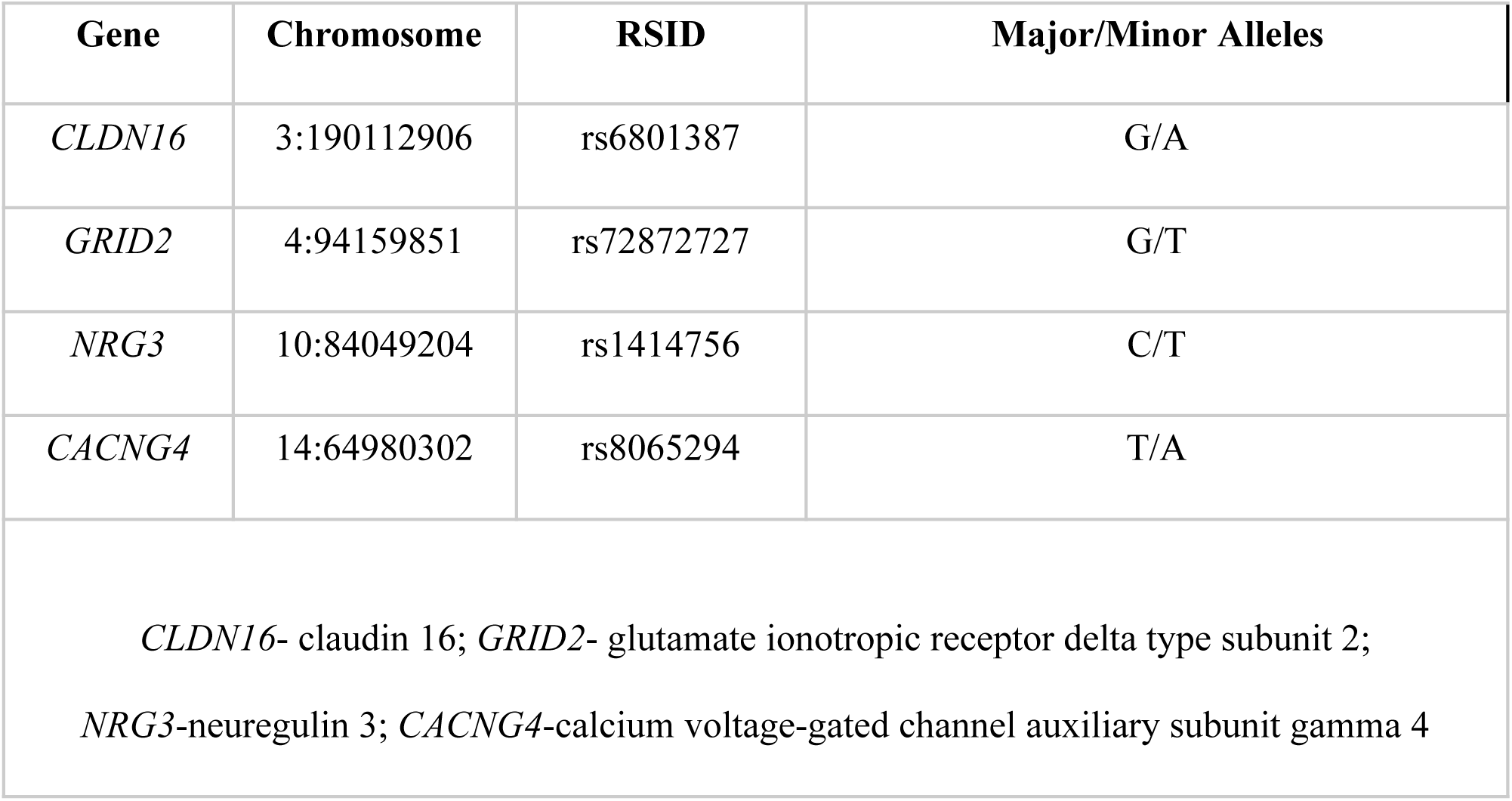
: Overview of the four single-nucleotide variations (SNVs)

| Gene | Chromosome | RSID | Major/Minor Alleles |
| --- | --- | --- | --- |
| *CLDN16* | 3:190112906 | rs6801387 | G/A |
| *GRID2* | 4:94159851 | rs72872727 | G/T |
| *NRG3* | 10:84049204 | rs1414756 | C/T |
| *CACNG4* | 14:64980302 | rs8065294 | T/A |
| *CLDN16*- claudin 16; *GRID2*- glutamate ionotropic receptor delta type subunit 2;<br>*NRG3*-neuregulin 3; *CACNG4*-calcium voltage-gated channel auxiliary subunit gamma 4 |  |  |  |

#### Overview of the four single-nucleotide variations (SNVs)

The evaluation of statistical differences commenced by examining the variations in age, glucose, and lipid metrics such as total cholesterol, LDL cholesterol, HDL cholesterol, triglycerides, and HbA1c between two groups. Given the data’s non-parametric characteristics, the Mann-Whitney U test (utilizing the Scipy library) was utilized for these analyses [30]. Gender distribution across the groups was evaluated using the Chi-square test, with detailed results presented in Table 2.

**Table 2.**
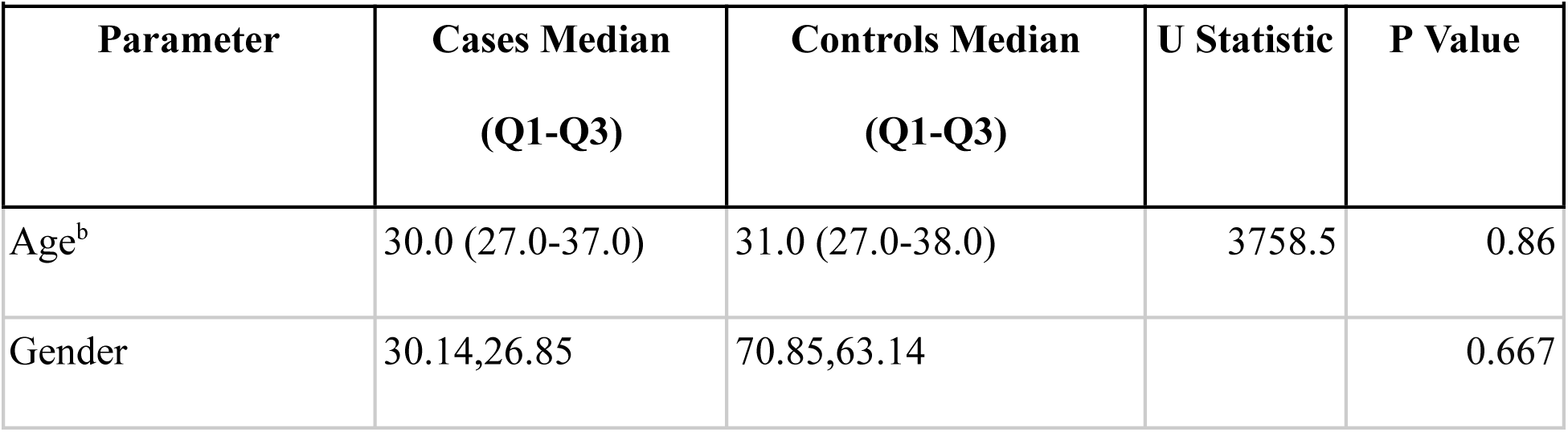
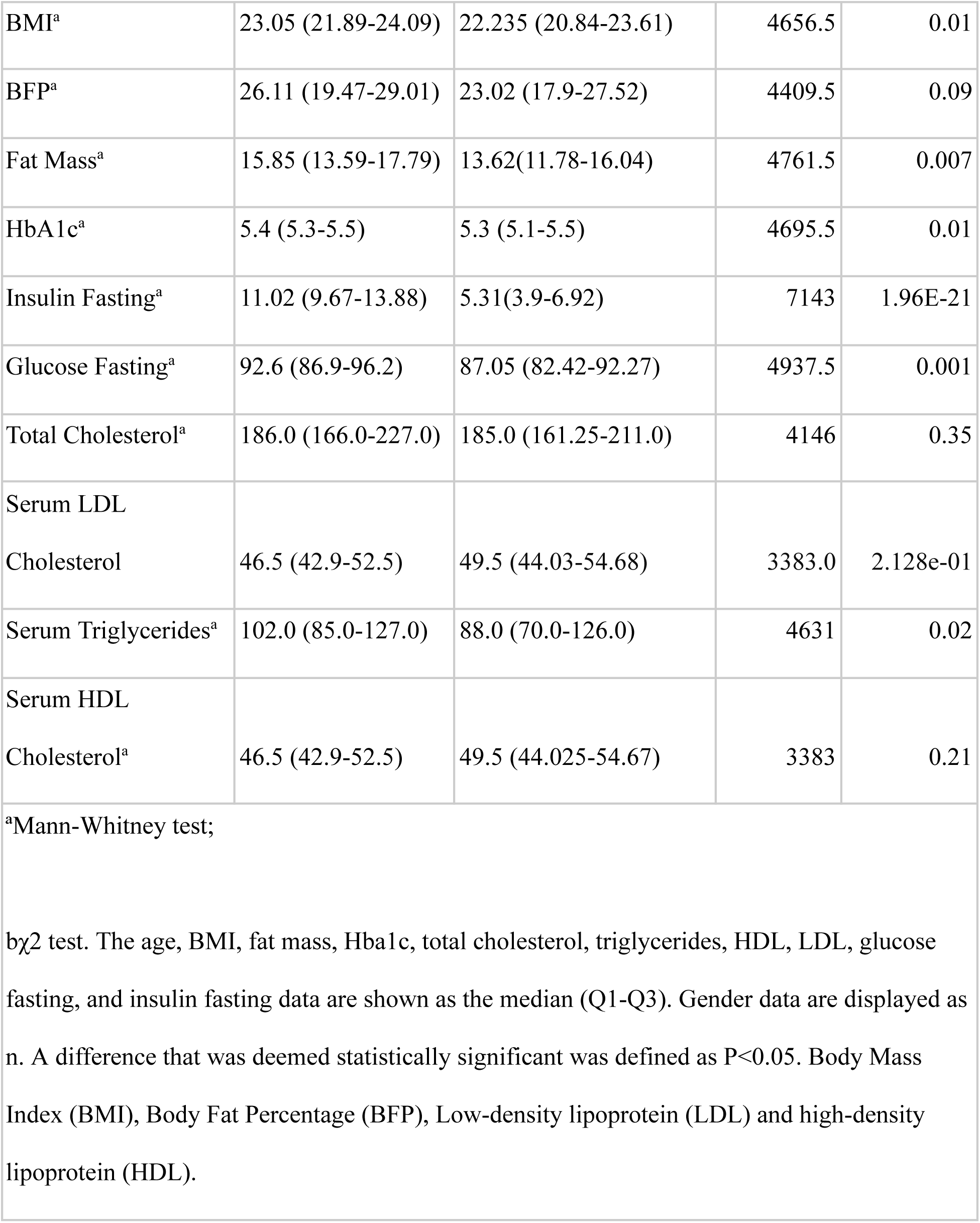
Characteristics of clinical significance and metabolic indicators related to glucose and lipid levels in participants of the current research.

### 4. Statistical analysis

Utilizing PLINK software [31] for quality control, the study filtered genotype data by removing samples with low genotyping rates, excluding SNPs with significant missing data, and eliminating rare variants for their minimal statistical power. Heterozygosity checks were also conducted to ensure accurate genotype distribution. Subsequently, a study was carried out using SNPstats software to ascertain the correlation between certain SNPs and IR. The analysis yielded odds ratios (ORs) together with their 95% confidence intervals (CIs). Table 3 summarizes the results, which show significant SNP markers under different genetic inheritance models.

**Table 3:** Comparison of the genotypic and allelic variations across four SNPs (rs6801387, rs72872727, rs1414756, rs8065294) between Cases and Controls.

| SNP | Group | A1 | A2 | $\chi^2$ | $\chi^2$<br>P-Value | A1A1 | A1A2 | A2A2 | P-Value | HWE<br>P-Value |
| --- | --- | --- | --- | --- | --- | --- | --- | --- | --- | --- |
| rs6801387 | cases | G<br>112<br>(0.98) | A<br>2<br>(0.02) | 12.67 | 0.00037 | G/G<br>55<br>(0.96) | G/A<br>2<br>(0.04) | A/A<br>0<br>(0) | 0.0005 | 1 |
|  | controls | G<br>231<br>(0.86) | A<br>37<br>(0.14) |  |  | G/G<br>99<br>(0.74) | G/A<br>33<br>(0.25) | A/A<br>2<br>(0.01) |  | 1 |
| rs7287272 | cases | G | T | 12.18 | 0.0004 | G/G | G/A | A/A | 0.0002 | 1 |
| 7 |  | 112<br>(0.98) | 2<br>(0.02) |  | 8 | 55<br>(0.96) | 2<br>(0.04) | 0<br>(0) |  |  |
|  | control<br>s | G<br>232<br>(0.87) | T<br>36<br>(0.13) |  |  | G/G<br>102<br>(0.76) | G/A<br>26<br>(0.21) | A/A<br>4<br>(0.03) |  | 0.26 |
| **rs1414756** | cases | C<br>96<br>(0.84) | T<br>18<br>(0.16) | 13.41 | 0.0002<br>5 | CC<br>40<br>(0.7) | C/T<br>16<br>(0.28) | TT<br>1<br>(0.02) | 0.0009 | 1 |
|  | control<br>s | C<br>176<br>(0.66) | T<br>92<br>(0.34) |  |  | CC<br>60<br>(0.45) | C/T<br>56<br>(0.42) | TT<br>18<br>(0.13) |  | 0.44 |
| **rs8065294** | cases | T<br>81<br>(0.71) | C<br>33<br>(0.29) | 14.41 | 0.0001<br>5 | CC<br>4<br>(0.07) | T/C<br>25<br>(0.44) | TT<br>28<br>(0.49) | 0.0006 | 0.75 |
|  | control<br>s | T<br>134<br>(0.5) | C<br>134<br>(0.5) |  |  | CC<br>37<br>(0.28) | T/C<br>60<br>(0.45) | TT<br>37<br>(0.28) |  | 0.23 |

Note: In the chi-square association analysis of cases and controls, the four SNPs (rs6801387, rs72872727, rs1414756, rs8065294) exhibited statistically significant associations with the trait after Bonferroni correction, with their respective p-values falling below the adjusted threshold of 0.0125.

In the control group, Hardy-Weinberg equilibrium was maintained with a significance level set at 0.05. The assessment of risk was conducted through the calculation of odds ratios (OR) along with their 95% confidence intervals (95% CI). The study also evaluated the linkage disequilibrium (LD) between the SNPs, with the LD coefficient D being determined via the use of Snpstats software [32]. Additionally, power analyses were conducted based on power and sample size calculations [33].Each SNP underwent a Bonferroni adjustment to account for the possibility of a type I error brought on by multiple tests.

## RESULTS

### Subject characteristics

In this study, all 191 participants were genotyped, with their clinical features and lipid metabolism indicators detailed in Table 2. The investigation revealed no meaningful statistical disparities in age or gender between the case and control groups, nor among males and females within those groups (p>0.05). Yet, notable distinctions were observed in BMI (0.01), Fat Mass (0.007), HbA1c (0.01), Glucose Fasting (0.001), and Serum Triglycerides (0.02), with the exceptions of BFP, Total Cholesterol, and Serum HDL Cholesterol when comparing cases to controls.

### Association of the 4 SNPs with IR

This study explored the relationship between four SNPs and insulin resistance in a normal-weight Indian population. We conducted a sample size calculation to ensure sufficient statistical power (power > 80%) to identify an effect size (OR) of greater than 1 for each SNP, considering a significance level of alpha = 0.05.

The frequency of genotypes for all four SNPs among both cases and controls was evaluated to examine their compliance with Hardy-Weinberg equilibrium (HWE) through a Fisher exact test. None of the SNPs demonstrated a significant deviation from HWE (p > 0.05), suggesting absence of genotyping errors and population stratification; the results are mentioned in Table 3.

We performed chi-square tests to evaluate the association between genotypes and allele frequencies of each SNP with phenotype (IR). After Bonferroni correction for multiple testing (p < 0.0125), all four SNPs displayed significant associations with IR:

The analyses were carried out between the genotypic and allelic variations of four SNPs (rs6801387, rs72872727, rs1414756, and rs8065294) within two groups distinguished by their HOMA2 IR levels (>2 and <2).The obtained p-values indicate significant associations between SNP genotypes and IR status.

- rs6801387 (*CLDN16*), the presence of the G allele was linked to a higher likelihood of insulin resistance (IR) (Odds Ratio (OR) = 9.18; 95% Confidence Interval (CI): 2.11-39.94; p < 0.001) when evaluated using the dominant model.
- rs72872727 (*GRID2*), carrying the T allele was found to elevate the risk of IR (OR = 11.14, 95% CI: 2.42-51.26, p < 0.0001) according to the dominant model.
- rs1414756 (*NRG3*), the T allele was identified as increasing the risk of IR (for C/T: OR = 2.38; 95% CI: 1.18-4.79; p = 0.0009; for T/T: OR = 12.2, 95% CI: 1.55-95.82, p = 0.0009).
- rs8065294 (*CACNG4*) demonstrated that the C allele significantly raises the risk of IR under the codominant model (for T/C: OR = 1.88; 95% CI: 0.94-3.74, p = 0.0006; for C/C: OR = 7.05, 95% CI: 2.23-22.23, p = 0.0006).

## MODEL OF INHERITANCE ANALYSIS

Tables 4 through 7 provide specifics on the genetic inheritance patterns (codominant, dominant, recessive, overdominant, and log-additive models) for these SNPs. Finding the lowest Akaike Information Criterion (AIC) and Bayesian Information Criterion (BIC) values was necessary to determine which model was best for each SNP.[32].

The optimal inheritance model for rs6801387 in *CLDN16* and rs72872727 in *GRID2*, was identified as dominant. For rs6801387, the risk genotype for IR was found to be (G/A-A/A) as opposed to the G/G genotype, with a significant association (P=0.0001, OR=9.18; 95% CI: 2.11-39.94) documented in Table 4. In the case of rs72872727, the risk genotype (G/T-T/T) was determined in contrast to the G/G genotype, showing a strong correlation (P<0.0001, OR=11.14; 95% CI:2.42-51.26) as shown in Table 5. For both rs1414756 in *NRG3* and rs8065294 in *CACNG4*, the codominant model was the most fitting. Regarding rs1414756, the risk genotypes (C/T and T/T) were identified against the C/C genotype for IR, with results (P=0.0009; OR= 2.38; 95% CI:1.18-4.79) and (P=0.0009; OR=12.20; 95% CI:1.55-95.82) presented in Table 6. For rs8065294, the (T/C) and (C/C) genotypes were marked as risk factors in comparison to the T/T genotype, with statistical significance (p= 0.0006; OR=1.88; 95% CI:0.94-3.74) and (p= 0.0006; OR=7.05; 95% CI:2.23-22.23) noted in Table 7.

**Table 4.** Different models of inheritance analysis of the SNP rs6801387 (*CLDN16*) between the Cases and Controls.

|  |  |  |  |  |  |  |  |
| --- | --- | --- | --- | --- | --- | --- | --- |
| rs6801387 association with response status (n=191, adjusted by gender+age.cat) |  |  |  |  |  |  |  |
| Model | Genotype | status=Ca | status=Co | OR (95% CI) | P-value | AIC | BIC |
| **Codominant** | G/G | 55 (96.5%) | 99 (73.9%) | 1 | 0.0005 | 231 | 260.3 |
|  | G/A | 2 (3.5%) | 33 (24.6%) | 8.63<br>(1.98-37.62) |  |  |  |
|  | A/A | 0 (0%) | 2 (1.5%) | NA (0.00-NA) |  |  |  |
| **Dominant** | G/G | 55 (96.5%) | 99 (73.9%) | 1 | **0.0001** | 229.2 | 255.3 |
|  | G/A-A/A | 2 (3.5%) | 35 (26.1%) | **9.18**<br>(2.11-39.94) |  |  |  |
| **Recessive** | G/G-G/A | 57 (100%) | 132 (98.5%) | 1 | 0.24 | 242.9 | 268.9 |
|  | A/A | 0 (0%) | 2 (1.5%) | NA (0.00-NA) |  |  |  |
| **Overdominant** | G/G-A/A | 55 (96.5%) | 101 (75.4%) | 1 | 0.0002 | 230.7 | 256.8 |
|  | G/A | 2 (3.5%) | 33 (24.6%) | 8.42 |  |  |  |
|  |  |  |  | (1.93-36.72) |  |  |  |
| **Log-additive** | --- | --- | --- | 8.76<br>(2.03-37.74) | 0.0001 | 229 | 255 |

**Table 5.** Analysis of various genetic inheritance patterns for SNP rs72872727 (GRID2) among case and control groups.

**rs72872727 association with response status (n=191, adjusted by gender+age.cat)**
| Model | Genotype | status=Ca | status=Co | OR (95% CI) | P-value | AIC | BIC |
| --- | --- | --- | --- | --- | --- | --- | --- |
|  | G/G | 55 (96.50%) | 102 (76.10%) | 1 |  |  |  |
| **Codominant** | G/T | 2 (3.50%) | 28 (20.90%) | 9.54<br>(2.08-43.74) | 0.0002 | 228.7 | 258 |
|  | T/T | 0 (0%) | 4 (3%) | NA (0.00-NA) |  |  |  |
| **Dominant** | G/G | 55 (96.50%) | 102 (76.10%) | 1 | **<0.0001** | 227.3 | 253.4 |
|  | G/T-T/T | 2 (3.50%) | 32 (23.90%) | **11.14**<br>(2.42-51.26) |  |  |  |
| **Recessive** | G/G-G/T | 57 (100%) | 130 (97%) | 1 | 0.065 | 240.9 | 266.9 |
|  | T/T | 0 (0%) | 4 (3%) | NA (0.00-NA) |  |  |  |
| **Overdominant** | G/G-T/T | 55 (96.50%) | 106 (79.10%) | 1 | 0.0003 | 231.1 | 257.1 |
|  | G/T | 2 (3.50%) | 28 (20.90%) | 8.67 |  |  |  |
|  |  |  |  | (1.94-38.81) |  |  |  |
| **Log-additive** | — | — | — | 9.94<br>(2.24-44.17) | <0.0001 | 226.8 | 252.8 |

**Table 6.** Different models of inheritance analysis of the SNP rs1414756 (*NRG3*) between the Cases and Controls.

**rs1414756 association with response status (n=191, adjusted by gender+age.cat)**
| Model | Genotype | status=Ca | status=Co | OR (95%CI) | P-value | AIC | BIC |
| --- | --- | --- | --- | --- | --- | --- | --- |
|  | C/C | 40 (70.2%) | 60 (44.8%) | 1 |  |  |  |
| ***Codominant*** | C/T | 16 (28.1%) | 56 (41.8%) | 2.38<br>(1.18-4.79) | **0.0009** | 232.3 | 261.6 |
|  | T/T | 1 (1.8%) | 18 (13.4%) | **12.20**<br>(1.55-95.82) |  |  |  |
|  | C/C | 40 (70.2%) | 60 (44.8%) | 1 |  | 233.8 | 259.8 |
| ***Dominant*** | C/T-T/T | 17 (29.8%) | 74 (55.2%) | 2.96<br>(1.50-5.82) | 0.0012 |  |  |
|  | C/C-C/T | 56 (98.2%) | 116 (86.6%) | 1 | 0.0052 | 236.5 | 262.5 |
| ***Recessive*** | T/T | 1 (1.8%) | 18 (13.4%) | 8.65<br>(1.12-66.72) |  | 236.5 | 262.5 |
|  | C/C-T/T | 41 (71.9%) | 78 (58.2%) | 1 |  |  |  |
| **Overdominant** | C/T | 16 (28.1%) | 56 (41.8%) | 1.85<br>(0.93-3.66) | 0.073 | 241.1 | 267.1 |
| **Log-additive** | — | — | — | 2.72<br>(1.53-4.86) | 0.0002 | 230.7 | 256.8 |

**Table 7.**
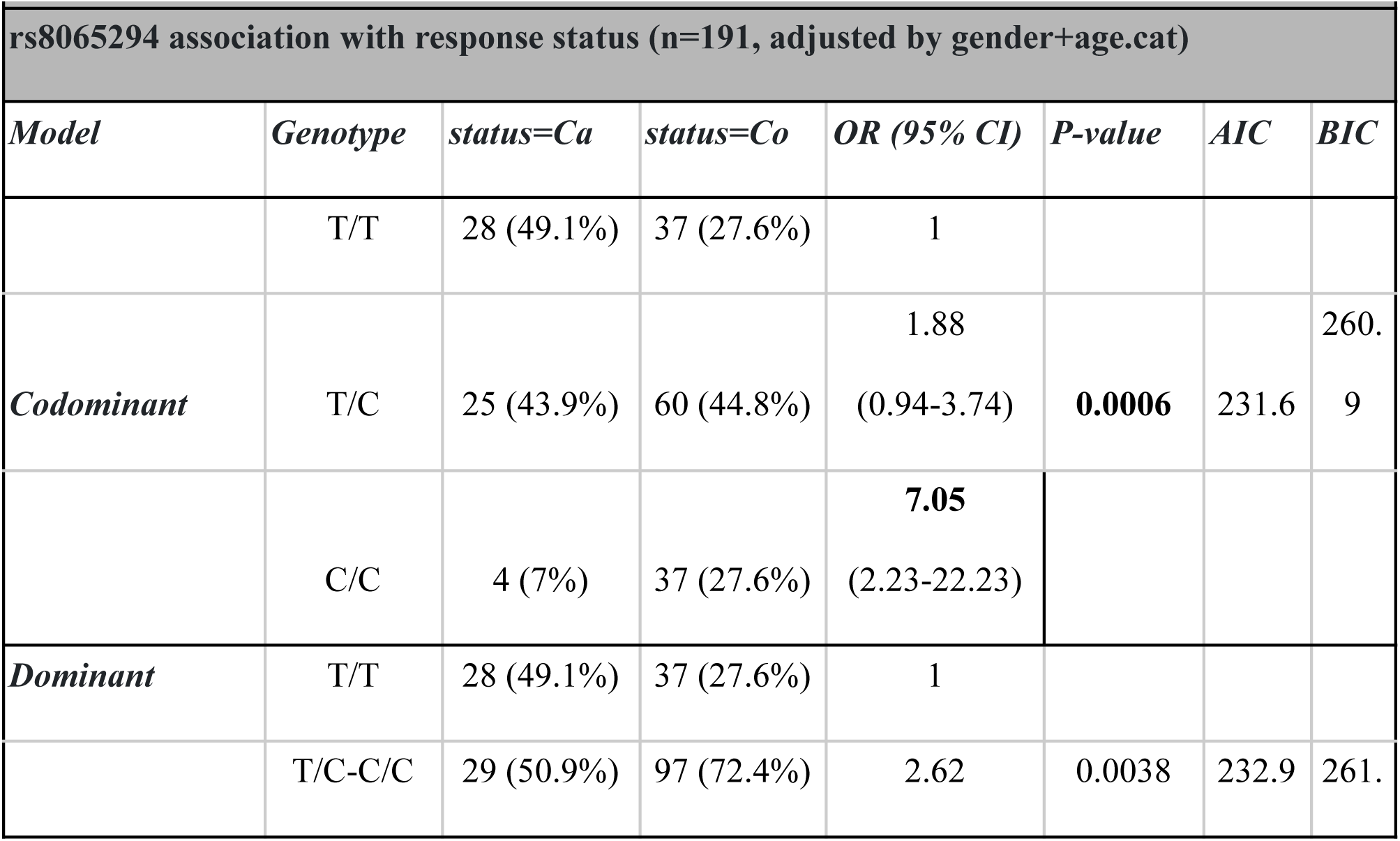
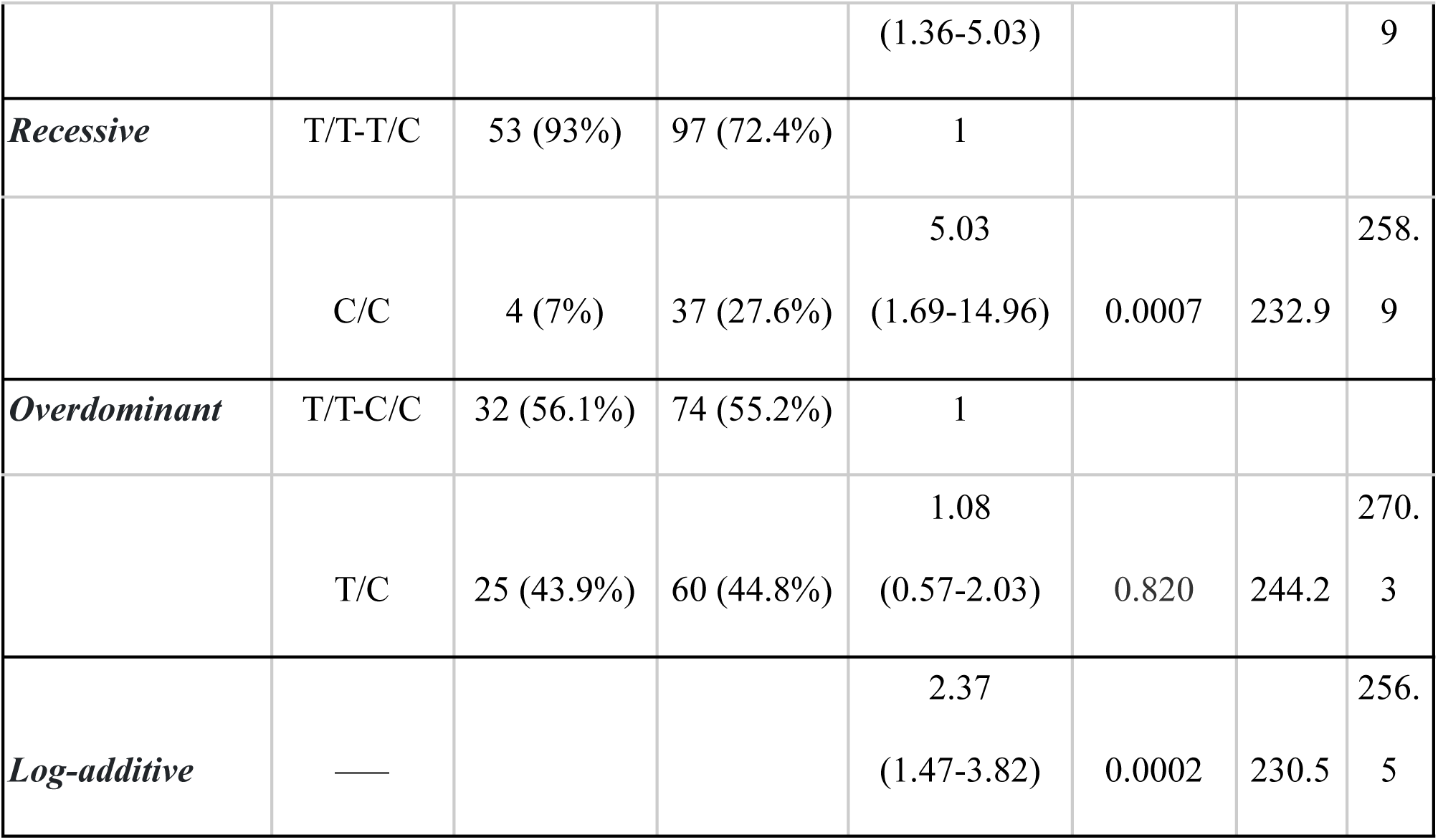
Different inheritance models analysis of the SNP rs8065294 (*CACNG4*) between the Cases and Controls.

These findings support the established role of these parameters in IR. It can be inferred that the development of IR in the normal-weight Indian population can be due to a combination of reported clinical factors and genetic abnormalities in certain genes including *CLDN16*, *GRID2*, *NRG3*, and *CACNG4*. To assess the underlying mechanisms and investigate possible treatment targets, more functional research is required.

## DISCUSSION

Claudin-16 (*CLDN16*) is distinctly expressed in the tight junctions of renal epithelial cells within Henle’s loop’s thick ascending limb, playing a pivotal role in divalent cation reabsorption. More than 20 mutations in the *CLDN16* have been discovered in individuals with familial hypomagnesemia characterized by abnormal renal excretion of Mg2+ and Ca2+. Prospective research on both genders has linked a higher intake of magnesium with a decreased risk of type 2 diabetes (T2D) [34] and metabolic disorders [35,36]. It was found that genetic polymorphisms associated with magnesium transport significantly elevate the risk of T2D in those with lower magnesium consumption, supporting the idea that Mg2+ transport alterations could hinder insulin response, thereby increasing T2D susceptibility [37, 38]. Chan et al identified a correlation between low magnesium intake and cellular Mg2+ imbalances with KATP channel mutations in influencing T2D risk [37]. Our findings demonstrate a link between the allele and genotypes of rs6801387 in the *CLDN16* and IR, with the A allele of rs6801387 identified as a significant risk factor for developing IR (P<0.001, OR=9.18; 95% CI: 2.11-39.94).

The human *GRID2* gene encodes a glutamate ionotropic receptor which is majorly expressed in the brain’s cerebellar Purkinje cells [39].The *GRID2* is located in a functional area that controls dietary consumption and energy balance.[40] In a murine research, the *GRID2* knockout mice’s gut microbiota showed great disruption in the neuroactive ligand-receptor activity. The *GRID2* could affect the levels of certain gut bacteria. The gut microbiome and interactions with brain and gut are set during pregnancy and early life, and can be affected by various factors like metabolism, diet, stress, and medications throughout life [41]. Any disruption in this BGM (Brain- gut- microbiome) system can lead to more pleasure-driven eating habits, cravings, and overeating leading to obesity [42,43]. The *GRID2* has been found within the locus affecting body weight in mice, and its connection to obesity in humans, particularly when combined with tobacco usage, as found in a family-oriented study [44]. Our present findings suggest a link between specific alleles and genotypes of rs72872727 in the *GRID2* and IR. The T allele of rs72872727 has been identified as a significant risk factor for IR, with statistical significance (P<0.0001) and an odds ratio of 11.14 (95% confidence interval: 2.42-51.26).

Neuregulin-3 (*NRG3*) has been identified as a novel protein that shares structural features with other neuregulins, especially neuregulin-1 (*NRG1*). *NRG1* and related neuregulins play key roles in the growth and cell differentiation. The action of *NRG1* is mediated through the activation of the *ErbB2*, *ErbB3*, and *ErbB4* receptor tyrosine kinases [45,46].Metabolic impacts of *NRG*, specifically regarding ErbB4 in skeletal muscle, have revealed that *ErbB4* levels are higher in skeletal muscle compared to liver, indicating potential metabolic implications.[46–50] Recent studies suggest that *ErbB4* polymorphisms are linked to metabolic complications indicating a major role of *Nrg4*/*ErbB4* signaling in its development [51–55]. Few studies have revealed a significant correlation between serum *ErbB2* levels and IR [56–59]. Our study’s results point to a relationship between IR and the genotypic and allelic profiles of the NRG3’s rs1414756. Particularly, it has been determined that the T allele of rs1414756 poses a risk for IR (p=0.0009, OR=12.2; CI:1.55-95.82).

Yang et al. have found that the secretion of insulin stimulated by glucose involves voltage-gated Ca2+ (CaV) channels, specifically highlighting the essential role of the gamma-4 subunit (CaVγ4) in maintaining the differentiated state of beta-cells. Various CaV channels, including alpha1, beta, alpha2delta, and gamma subunits are expressed in beta-cells [60]. Wu R et al. ’s work with a murine model identifies *CACNG4* as vital for maintaining normal glucose homeostasis, highlighting CaVγ4 as a potential target for prediabetes treatment by amending impaired metabolic conditions [61]. Additionally, Luan C’s research underscores the importance of CaVγ4 in sustaining the functionality and differentiated state of beta-cells, suggesting that treatments targeting CaVγ4 could aid in restoring beta-cell functionality in diabetic conditions [62]. Our results also show a connection between the allelic and genotypic variations of rs8065294 within the *CACNG4* and IR, highlighting the C allele of rs8065294 as a notable risk factor for the onset of IR (p=0.0006, OR=7.05; CI:2.23-22.23).

## CONCLUSION

In our current study, we carried out association studies on SNPs of four genes (*CLDN16*, *GRID2*, *NRG3*, and *CACNG4*) and IR among the normal BMI Indian population. We selected these SNPs, as they have some literature presence involving IR. Our study revealed that the individuals carrying the allele G for rs6801387, allele T for rs72872727 and rs1414756 and allele C for rs8065294 are at higher risk for developing IR. Also, The SNP rs72872727 in *GRID2* (p<0.0001;OR=11.14; 95% CI(2.42-51.26) has the greatest significance, beyond the genetic factors, certain clinical parameters such as Fat Mass, Serum Triglycerides, Hba1c have shown significant differences between cases and controls, suggesting a multifactorial etiological of the condition under study.

Although these genetic indicators demonstrate a significant predisposition towards the conditions under investigation, it’s crucial to consider the constraints of this study, such as the relatively small sample size that could affect its statistical robustness, underscoring the necessity for additional verification across broader and more varied cohorts.The future research could explore the functional significance of these SNPs and its potential mechanisms underlying the response trait.

## Abbreviations

(IR): Insulin resistance
(SNPs): single nucleotide polymorphisms
(HOMA2-IR): Homeostasis Model Assessment for Insulin Resistance
(GSA): Global Screening Array
(DM): Diabetes Mellitus
(NCDs): noncommunicable diseases
(BMI): Body mass index
(BF%): body fat percentage
(TG): triglycerides
(LDL-C): high-density lipoprotein cholesterol
(LDL-C): low-density lipoprotein cholesterol
(TC): total cholesterol
(FPG): and fasting plasma glucose
(ORs): odds ratios
(CIs): confidence intervals
(LD): linkage disequilibrium
(HWE): Hardy-Weinberg equilibrium
(AIC): Akaike Information Criterion
(BIC): Bayesian Information Criterion
(T2D): type 2 diabetes

## Data Availability

All data produced in the present study are available upon reasonable request to the authors

https://www.nugenomics.in/

## Declarations

We confirm that this work is original and has not been published elsewhere, nor is it currently under consideration for publication elsewhere.

## Ethics approval and consent to participate

This study was approved by the Answergenomics Ethical Review Committee Board. We have obtained written consent from all the subjects.

## Availability of data and materials

The datasets generated and/or analyzed during the current study are not publicly available due to the sensitive nature of genetic data and privacy concerns for participants. However, data are available from the corresponding author upon reasonable request and with the assurance that the privacy of participants will be maintained.

